# Impact of sample size and tissue relevance on T2D gene identification

**DOI:** 10.1101/2024.10.31.24316435

**Authors:** D Davtian, T Dupuis, D Mansour Aly, N Atabaki-Pasdar, M Walker, P W Franks, F Rutters, HK Im, E R Pearson, M van de Bunt, A Viñuela, AA Brown

## Abstract

Identification of genes and proteins mediating the activity of GWAS variants requires molecular data from disease relevant tissues, but these may be difficult to collect. Using multiple gene expression reference datasets and GWAS summary statistics for T2D we identified 1,818 unique genes associated with T2D. Comparing the performance of different reference datasets, we found that sample size, and not the relevance of the tissue to the disease, was the critical factor in identifying relevant genes. Genes implicated using a well powered expression dataset were also more likely to have multiple lines of genetic evidence. A targeted proteomics reference dataset from plasma samples showed similar power to identify T2D related proteins as gene expression with the same sample size. Accounting for BMI reduces power across all tissues and phenotypes by ∼30%, suggesting that many GWAS links to T2D are mediated by BMI, potentially implicating insulin resistance related effects. Finally, using data from smaller GWAS studies with precisely defined T2D subtypes uncovers genes directly relevant to that subtype, such as *LST1*, an immune response gene for Severe Autoimmune Diabetes and *TRMT2A*, involved in beta-cell apoptosis, for Severe Insulin Deficient Diabetes. Our work demonstrates the benefits of well powered reference datasets in accessible tissues and well-defined disease subtypes when studying complex diseases involving multiple tissues.

## Introduction

Genome wide association studies (GWAS) in large cohorts have greatly expanded our knowledge on the genetic causes of complex diseases such as T2D, implicating hundreds of novel loci^1–3^. However, most of these genetic loci are located in non-coding regions of the genome. To understand the immediate consequences and function of non-coding variation, molecular studies have assayed gene expression in multiple human tissues and linked genetic variation to changes in gene expression. These studies provide a path for discovering disease causing genes and the tissues in which they are active by connecting GWAS loci to genes in a particular tissue^4–6^. Transcriptome wide association methods are a type of method using reference expression datasets to build predictive models of genetically regulated gene expression. These models are then applied to GWAS studies to produce cis-predicted expression phenotypes across thousands of genes, which can be tested for association with the disease. Where individual level data is unavailable, approximations exist using summary statistics. It has been argued that TWAS methods discover genes that are causal of the disease, because these are based solely on genotype data that predates the development of disease. However, this interpretation is complicated by issues such as pleiotropy (SNPs associated to the expression of several genes) and linkage contamination (correlated SNPs where one affects disease risk and the other expression)^7^. This means that while the methodology may implicate a particular gene, the true causal drivers may be other variants and genes nearby. For this reason we will refer to TWAS significant associations as causal loci rather than causal genes.

Reference datasets used to predict gene expression should be based on tissues that are directly relevant for the disease, to ensure that the tissue specific genetic effects that alter disease risk are captured^8^. However, many disease relevant tissues are difficult to collect, limiting the sample size of such experiments and the ability of TWAS methods to produce accurate predictors of gene expression (Figure 1A). Studies with small sample size are best powered to discover expression quantitative trait loci (eQTLs) with the largest effect size, these are typically located in promoter regions^9^ and have cross tissue effects, reducing the identification of tissue specific genetic effects. For example, Type 2 diabetes (T2D), a complex disease characterized by increased levels of glucose in blood^10^, involves multiple tissues with physiological and genetic factors influencing its pathogenesis, such as insulin resistance, obesity and beta-cell dysfunction. For each mechanism of action, different tissues and organs play greater and lesser roles. Insulin resistance related T2D is often associated with molecular changes in skeletal muscle, adipose and liver, while beta-cell dysfunction is almost exclusively associated with the pancreatic islets, where beta-cells are localized ^11^. Many of these relevant tissues are difficult to collect pre-mortem, with consequences for our ability to discover the genetic and molecular processes involved in the development of T2D. On the other hand, gene expression studies with thousands of samples have shown that the expression of most genes is affected by multiple eQTLs and many of these effects are shared across multiple tissues^12,13^. Therefore, it is possible that larger molecular studies using samples from tissues that are easy to collect, such as blood, could identify disease relevant loci through these shared signals.

**Figure 1.**
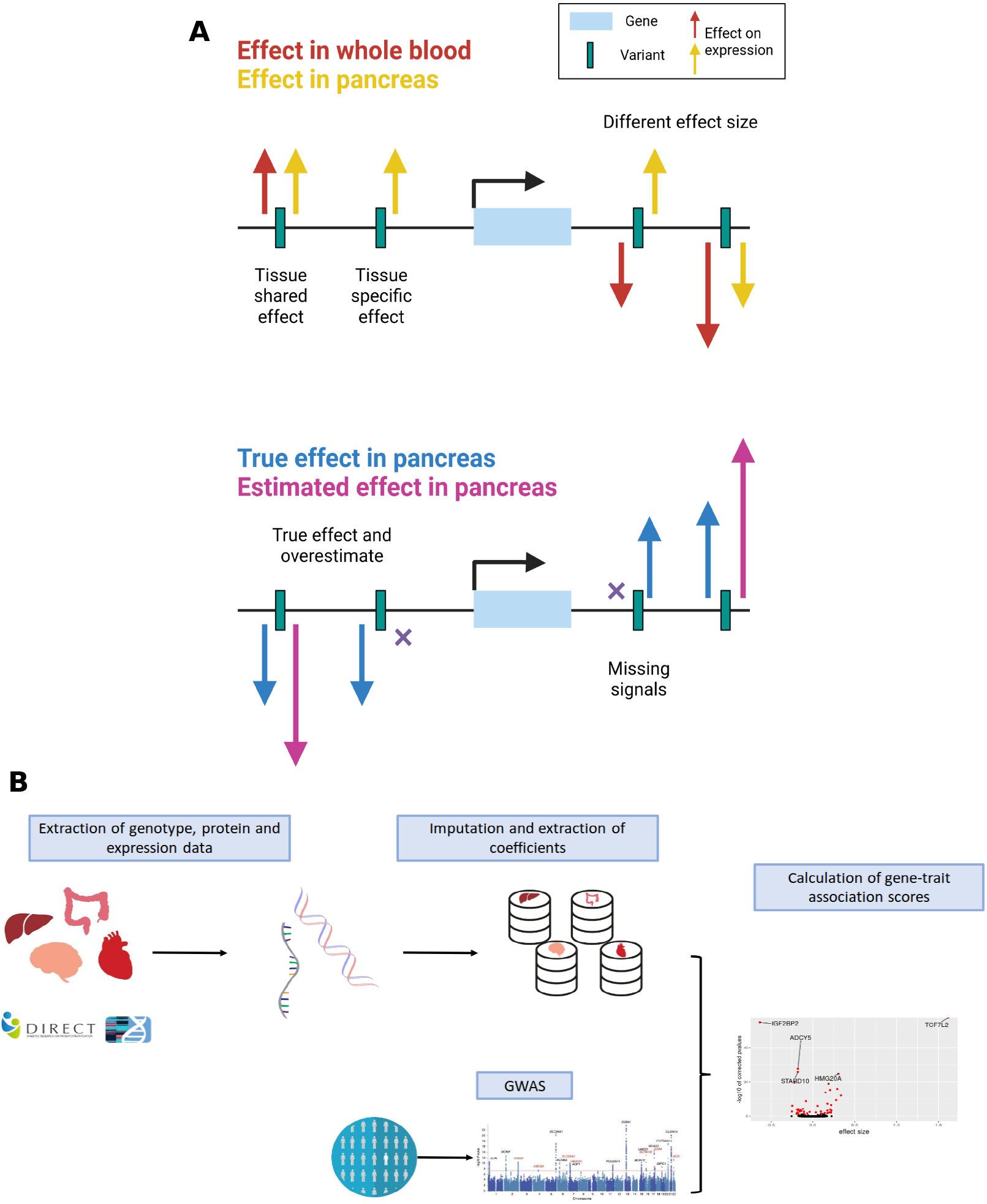
A. Schematic overview of the tradeoff between tissue specificity and sample size and their impact on detection of effects on expression. The upper section shows the consequences of using different tissues on the effect captured with TWAS. The lower section shows the different scenarios when the sample size is small. **B. Study workflow**. We developed prediction models for genetically regulated gene expression and protein levels in multiple datasets with different sample sizes. Predictive models were then associated to T2D using GWAS summary statistics to identify differentially expressed genes with T2D.

Here we aim to identify T2D relevant genes using TWAS methods in a large dataset from whole blood, an accessible tissue. We assessed the roles played by sample size and tissue relevance when identifying disease relevant genes using TWAS methods. We compared the ability to find T2D related genes using a large whole blood expression dataset produced by the DIRECT consortium (n=3,029) to the performance using relevant tissues but with smaller sample size (GTEx consortium, n<706, and pancreatic islets from InsPIRE, n=420). We show that sample size is the chief determinant of discovery power for associations between gene expression and disease risk. Furthermore, we explored the effect of considering environmental factors such as BMI on discovering T2D loci and the use protein-QTL (pQTL) data to gain potential biological insights on the disease. Finally, using GWAS data from T2D subtypes, we identify novel loci implicated in Severe Insulin Deficient Diabetes.

## Results

To identify T2D causal loci we used gene expression reference datasets produced by three consortia: DIRECT (whole blood, sample size 3,029), 49 tissues from GTEx (sample size 70 to 706) and InsPIRE (pancreatic islets, 420) (Supplementary table 1). These datasets were chosen to represent a full spectrum of modestly sized expression references generated from tissues that are directly relevant to T2D (pancreatic islets, subcutaneous and visceral adipose tissue, liver, skeletal muscle and kidney cortex) to a large dataset generated from a non-relevant tissue (whole blood, DIRECT)^4,12,14^ (Methods). Transcriptome wide association studies construct models of gene expression from reference expression datasets and use these to predict expression in GWAS studies, which can then be used to test for association with disease. In this study, expression was modeled using conditionally independent cis-eQTLs released by each consortium (Supplementary Table 1) and then associated with T2D using GWAS summary statistics from the DIAGRAM consortium and the MetaXcan method (Figure 1B)^15^. By adopting this approach the results were directly comparable across studies, as all studies used the same pipeline to generate conditionally independent cis eQTLs.

We identified 404 T2D associated loci using the whole blood DIRECT data (n=3,029 samples). This is the largest number of significant loci discovered across the reference panels, with 108 loci identified using the pancreatic islet InsPIRE dataset (n=420) and from 3 to 299 loci depending on tissue for GTEx (n<706) (1,568 in total) (Figure 2A – C, Supplementary table 2). We found a strong linear correlation between the sample size of the dataset and the number of significant loci discovered (Spearman correlation 0.95, p=7.62e^-27^, Figure 2B). However, the DIRECT dataset did not follow the linear trend, identifying fewer loci than would be predicted (Figure 2B). This could be due to limitations specific to the DIRECT data, such as lower sequencing depth and multi-centre sample collection, or due to a saturation of signals to discover disease loci^14^. We also tested for an effect of tissue relevance, using a linear model excluding DIRECT of discovered loci against sample size and found that fewer loci were discovered using relevant tissues than would be expected based on sample size (estimate = -29.27, p-value = 0.019). In summary, we found a linear relationship between sample size and discovery of disease relevant causal loci with little influence of the relevance of the tissue for the disease.

**Figure 2.**
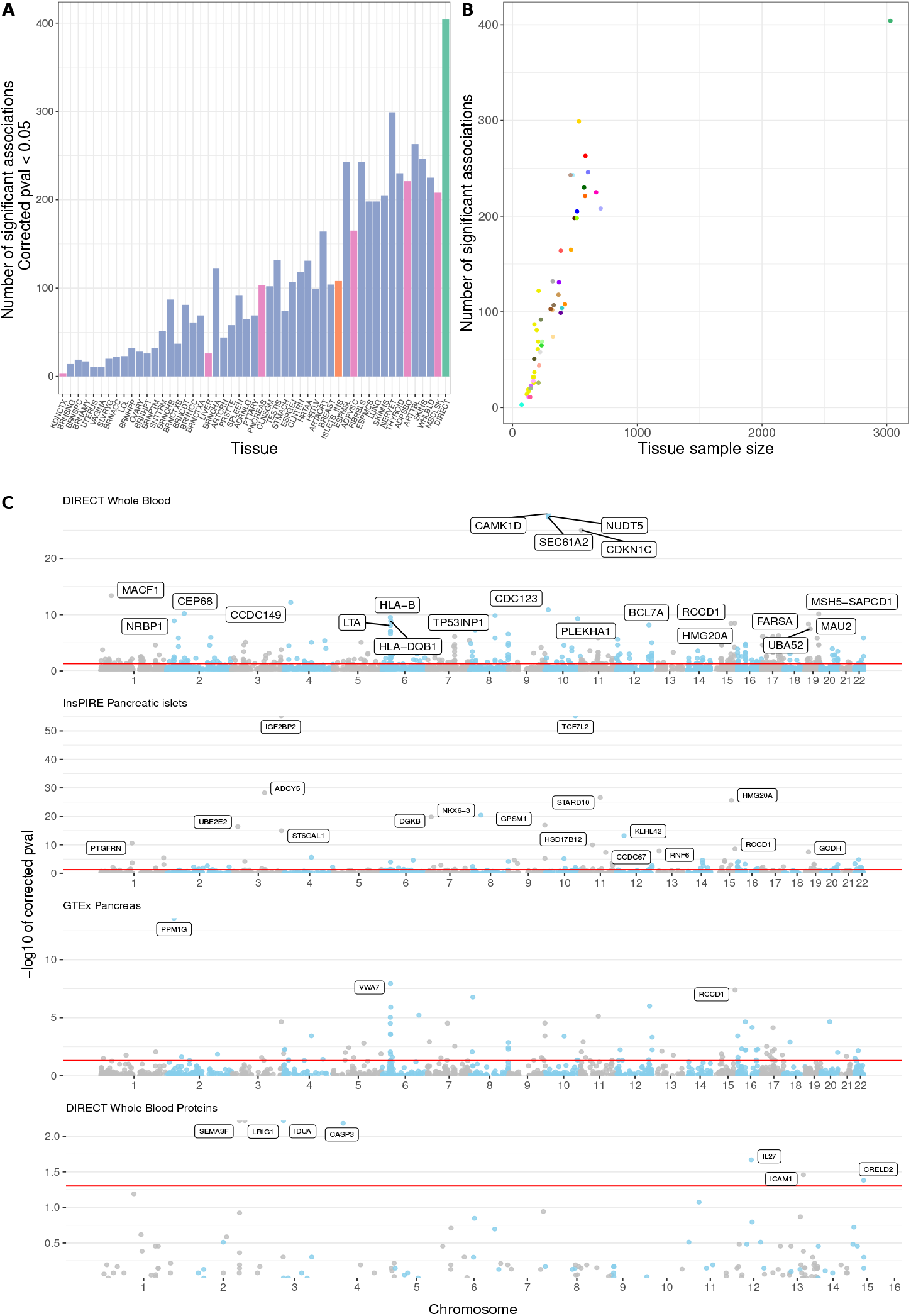
A. Distribution of the number of significant associations obtained with MetaXcan for each tissue tested. Tissues are ranked by sample size and colored by datasets or disease relevance, green is DIRECT, orange is InsPIRE, pink and purple GTEx with pink showing relevance for T2D. **B. Scatter plot showing *relationship* between sample size and number of significant associations**. Colors use the GTEx official color palette plus DIRECT in green and InsPIRE in orange. **C. MetaXcan results for three tissues and DIRECT proteins in a Manhattan style**. The red line indicates the significance threshold at –log10 (0.05). The labelled genes or proteins, indicate highly significant associations (p < 5e-8 for genes and p< 0.05 for proteins).

Large reference datasets identified more disease loci than smaller datasets, but the proportion of already known T2D loci was larger for relevant tissues (figure 3A). DIRECT identified 88 out of 404 (21.8%) of significant associations that were previously reported as related to T2D^16^. Across all GTEx tissues there were 251 known T2D loci out of a total 1,568 (16%, from 0 to 56 per tissue, Supplementary Table 3). In comparison, 30 out of 108 (28%) of the islets associated loci had previously been associated with T2D. For example, *UBA52* and *NAGLU* were among the DIRECT reported genes, which have previously observed to be over expressed in pancreas of individuals with T2D^17^. However, only using pancreatic islets data were we able to identify an association between T2D and *TCF7L2*, a gene located near one of the strongest known T2D GWAS signals^18^. Overall, we identified 1,515 potentially novel T2D genes across all tissues.

**Figure 3.**
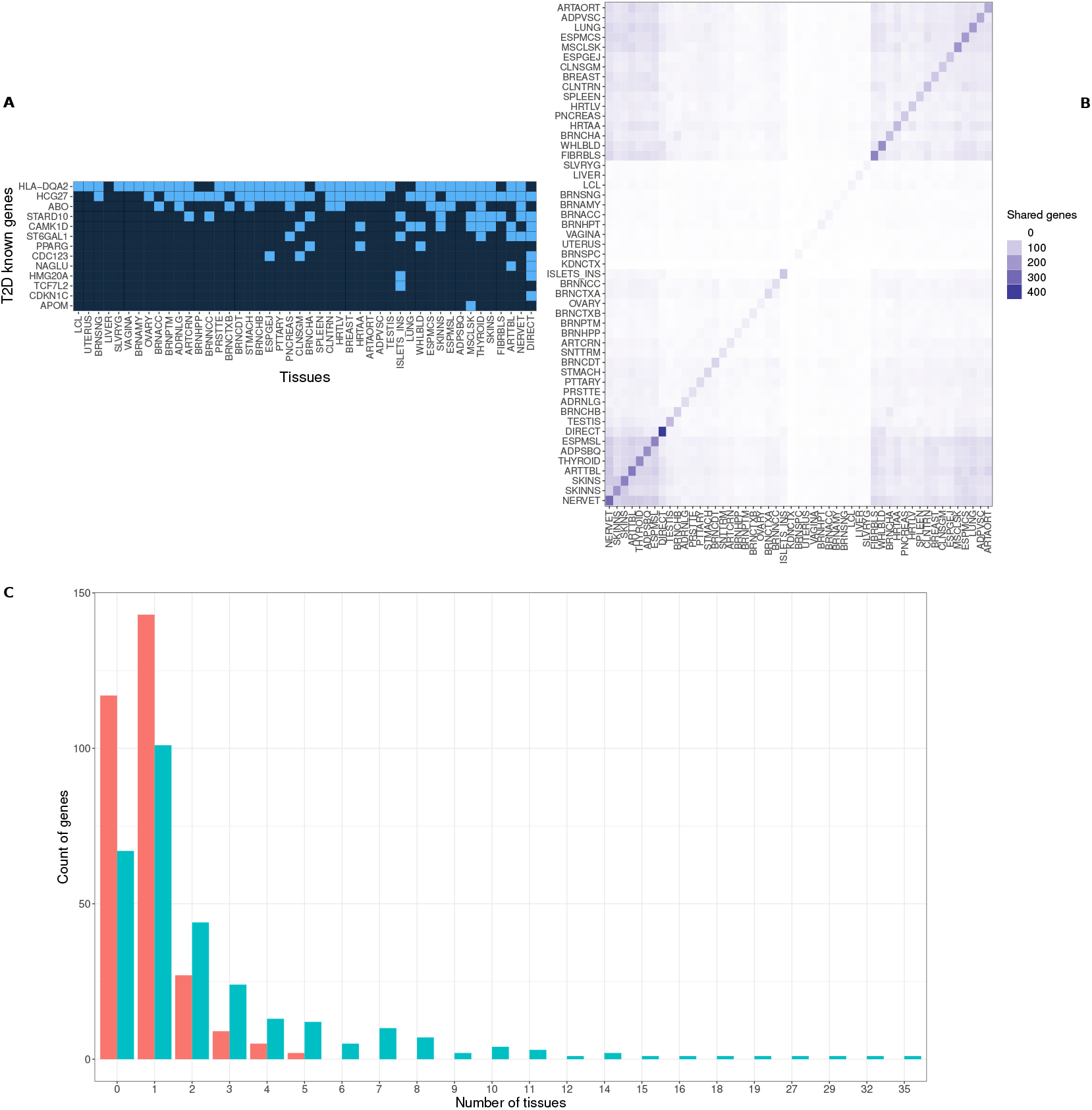
A. Heatmap of occurrences of 13 known T2D genes in all significant associations. Light blue tiles designate the presence of the gene and dark blue the absence. **B. Heatmap of the number of shared significant association between each pair of tissues. C. Barplots representing the distribution of number of occurrences of significant known T2D genes in all tissues separated by the relevance of the tissue**. Red bars are for counts in relevant tissues and green bars are counts in non-relevant tissues.

We next investigated the degree to which the reported genes were discovered in multiple tissues. Comparing the genes reported in significant loci across datasets, we observed that more than half of all significant genes were only discovered in one tissue: 196 were identified using only blood (49% of all DIRECT significant associations) while 51 were found using only pancreatic islets (47% of InsPIRE associations) (Supplementary table 4). Across tissues, we observed that similar tissues did not show any patterns of sharing genes (Figure 3B). This is consistent with most associations being driven by cross tissue effects, with a significant “winner’s curse” contribution to the discovery set in each tissue (Figure 3B). Moreover, we found that disease relevant tissues such as islets, liver and muscle, display more unique well known T2D genes compared to non-relevant tissues (Figure 3C). However, the pancreatic islets dataset missed well known T2D genes such as *CDKN1C*, involved in beta-cell proliferation and insulin responsiveness. This gene was only discovered using the increased power of the DIRECT reference. Our results show that studies with larger sample sizes such as DIRECT and the combined data of GTEx tissues, were able to identify more potentially novel genes (respectively 316 and 1,317) compared to the relevant pancreatic islets tissue (78). Moreover, we show that processes in non-relevant tissues may be informative of genetic effects relevant to disease, identifying novel disease candidate genes that would be missed due to sample size limitations in tissues considered relevant to the disease.

To better understand the nature of the SNPs underlying significant associations, we evaluated how well particular reference panels corroborated the loci implicated by two GWAS studies and investigated whether loci associations could be driven by individual, highly significant signals. We used the 492 T2D GWAS significant loci reported by Mahajan et al^19^ (the GWAS used in the TWAS analysis), and the 186 signals reported by Spracklen et al^20^, which used populations from a different ethnicity. We found that DIRECT associated loci were more likely to be within 1MB of a significant GWAS signal than all loci from each of the GTEx tissues (Figure 4A). Compared to islets loci, the DIRECT loci were less likely to be found near the GWAS signals of both studies (OR_Mahajan_ = 1.16 and OR_Spracklen_ = 1.22, Supplementary table 5). This enrichment of DIRECT and InsPIRE loci relative to GTEx may be a result of loss of information when lifting over GWAS summary statistics to GRCh38 genome build, necessary to combine these statistics with GTEx reference panels. However, when considering GTEx tissues alone, the relevance of the tissue for T2D did not predict enrichment, with tissues such as adipose subcutaneous and pancreas depleted relative to the median tissue (Figure 4A). There was also no correlation between sample size and the enrichment of significant genes around GWAS loci (Figure 4B).

**Figure 4.**
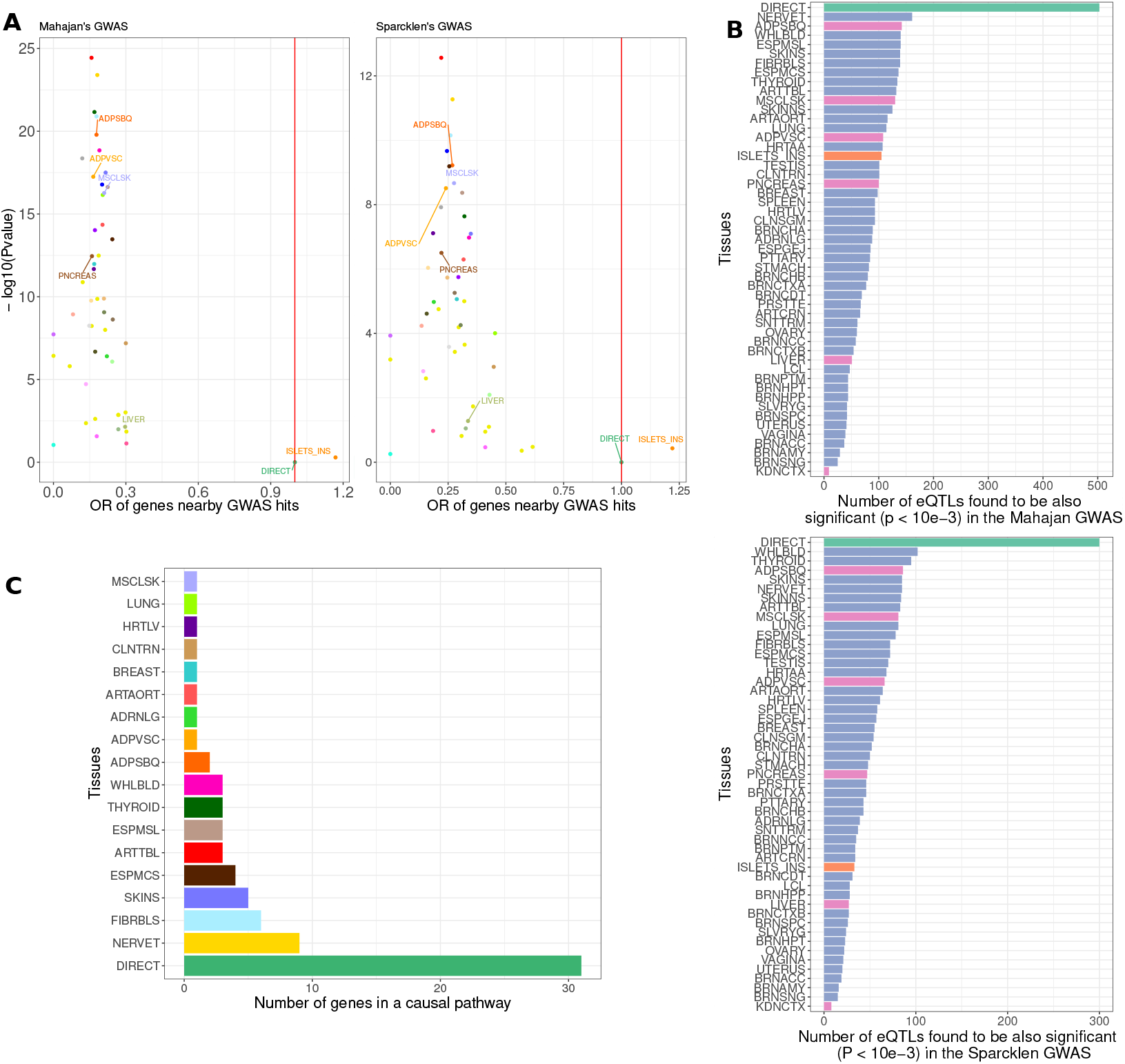
A. Proportion of significant genes located nearby significant GWAS hits from different studies. Odds ratio have been calculated relative to DIRECT findings allowing to display on the right side of the red line tissues getting better results than DIRECT and on the left side of the line the opposite. **B. Barplot of tissues ranked relative to their number of eQTLs found to be significant in the two GWAS (P < 10e-3)**. Colors are green for DIRECT, orange for InsPIRE, pink and purple GTEx with pink showing relevance for T2D. **C. MR analysis results**. We show the number of genes involved in causal pathways for each tissue tested.

We hypothesized that the increase in the number of discovered loci by the DIRECT dataset, could be driven by loci involving multiple expression variants with moderate effects on T2D risk that are hard to find in smaller datasets. The DIRECT study originally reported more than 60,000 independent cis-eQTLs, while in this study we found that more DIRECT eQTL show suggestive association with T2D risk GWAS results: 503 eQTLs for DIRECT had P<0.001 in the Mahajan GWAS, compared to 105 eQTLs for islets (Figure 4B, Supplementary table 6). To test more formally that more DIRECT loci were identified due to multiple eQTL effects on T2D risk, we applied multiple instrument Mendelian Randomization (MR). As proposed by the SMR and HEIDI methods^21^, we tested whether eQTL variants have consistent, non-zero effects on T2D risk. Using a Wald Ratio test, we looked for evidence of genetic effects affecting both traits, and applied a test of heterogeneity to evaluate the consistency of effects. We confirmed 31/214 of the loci with this approach in DIRECT (corrected pvalue < 0.05 and a heterogeneity test pvalue > 0.05), compared to between 1 and 9 across the other reference datasets. Therefore, we conclude that well powered reference datasets identified more loci with multiple variants with consistent effects on T2D risk than smaller reference panels (Figure 4C, Supplementary table 7). These loci were supported by a larger number of SNPs located further away from the target genes, and therefore with moderate genetic effects, compared to SNPs used from smaller studies.

While genetic effects on disease risk are expected to be mediated by transcription at some point, for many diseases protein levels have been shown to be more informative biomarkers^22,23^. We investigated if TWAS methods could identify T2D mediating protein loci with similar properties as gene expression by constructing models based on *cis* plasma targeted protein-QTLs (pQTLs). Using DIRECT genetic data and pQTLs for 149 proteins, we identified 7 loci associated with T2D:SEMA3F, IDUA, LRIG1, CASP3, IL27, ICAM1 and CRELD2 (Figure 2C). Among these, LRIG1, ICAM1 and CASP3 were coded by known T2D genes: LRIG1 is part of a protein family which is involved in lipid homeostasis and has been associated with T2D ^24^, ICAM1 has been shown to be involved in diabetic retinopathy ^25^ and expression of *CASP3* has been associated with islet apoptosis ^26,27^. Of the significant proteins, only LRIG1 and ICAM1 reported significant associations also with the genetically predicted gene expression for the coding gene, and for LRIG1 the reported associations showed opposite direction of effect (Zscore_expression_ = -3.43, pvalue_expression_ = 0.013, Zscore_protein_ = 3.85, pvalue_protein_ = 0.006). Despite the smaller number of loci identified with protein data, the proportion of associated loci *vs*. tested loci was approximately the same for proteins as it was for genes (4.6% compared to 4.7%). This suggests that both assays had similar power to discover mediating factors, though the genome-wide ability of RNA-seq to quantify mRNA means that more loci were tested and therefore discovered. Finally, we looked to see if for significantly associated proteins we were also more likely to find a significant association between disease and the corresponding gene, and we observe a large odds ratio for enrichment, though this was not significant due to the small number of proteins associated (odds ratio = 9.56, Fisher’s Exact test pvalue = 0.51). This large odds ratio is consistent with other studies, that have found that genetic effects on proteins tend to replicate on expression^13^.

T2D is a heterogeneous disease, involving different clinical presentations that go beyond the classical categorization of T2D as driven by beta-cell dysfunction or insulin resistance. Previous studies have shown that processes occurring in the pancreatic islets are likely related to T2D by beta-cell dysfunction, while obesity related T2D or insulin resistance are linked to expression patterns in adipose or muscle tissues^4,28^. However, T2D GWAS studies have shown that most loci do not change their effect size when controlling for BMI^20^. To explore disease subtypes causal loci, we repeated our association analysis using T2D GWAS summary statistics which controlled for BMI. We found a reduction of 732 genes discovered overall, from 1,818 unique genes to 1,086 and a fall in the number of protein associations from 7 to 4. This reduction we conjectured was due to the removal variation related to an important environmental predictor for T2D and those loci whose effect was mediated by BMI and thus more likely to relate to insulin resistance (Figure 5A-B, Supplementary table 8). However, the reduction impacted all tissues to a similar degree, with no greater depletion for adipose relative to islets tissue, suggesting that either beta-cell dysfunction effects are also partly related to obesity and active in multiple tissues or that we lack the ability to identify tissue or disease specific sub-types and processes. The reduction is also likely partly due to the reduced sample size of the GWAS controlling for BMI.

**Figure 5.**
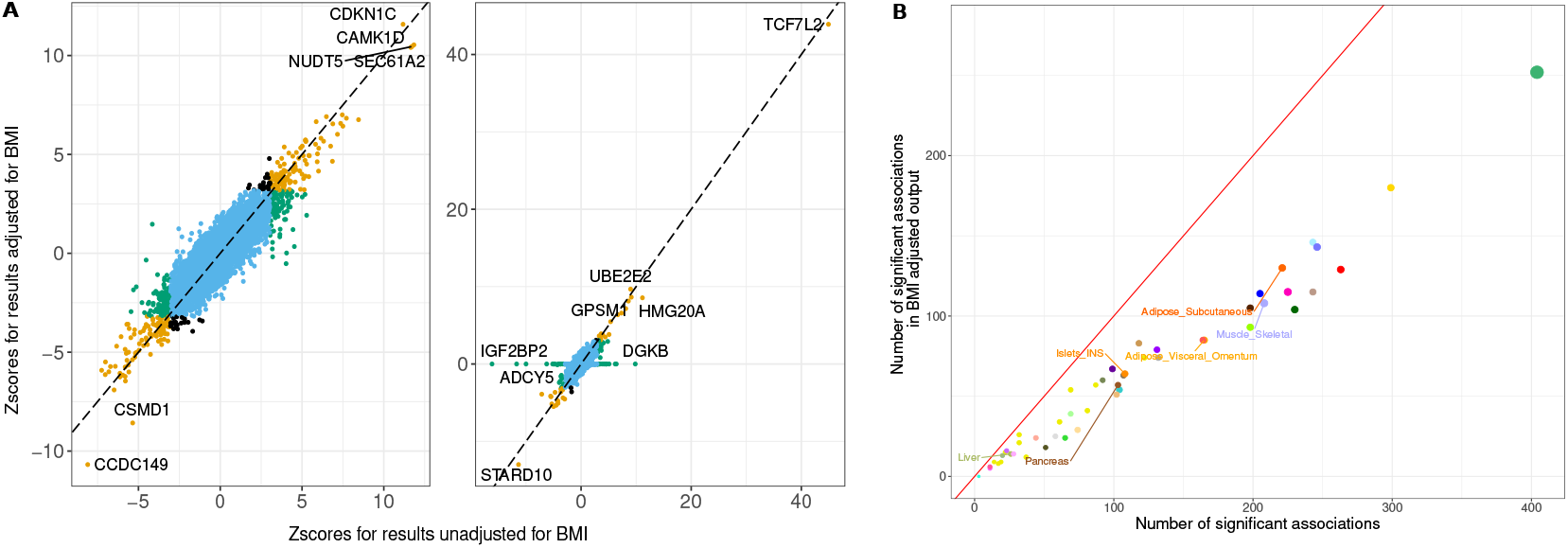
A. Comparison between results obtained with MetaXcan when adjusting for BMI or not. Are shown, Zscores for DIRECT and *InsPIRE*. The colors are blue for non-significant associations in both outputs, orange for both, green for only unadjusted results and black for adjusted results only. **B. Comparison of the overall number of significant associations for each tissue when controlling for BMI**.

Give the heterogeneity of T2D, a more specific definition of cases in genetic studies by their disease subtype can allow the discovery of disease loci specific to particular aspects of disease. Recently, Ahlqvist, et al. suggested that T2D could be considered as five distinct disease subtypes, namely severe autoimmune diabetes (SAID), severe insulin deficient (SIDD), severe insulin resistant diabetes (SIRD), mild obesity related (MOD) and mild age related diabetes (MARD)^29^. A GWAS study, considering these five subtypes separately, found genetic signals specific to particular subtypes and not discovered using larger GWAS studies into T2D, despite the smaller sample size for the disease subtypes GWAS (10,927 patients compared to ∼1,000,000)^30^. Applying our TWAS framework to their GWAS, we found that the size of the GWAS study also had an impact on the number of loci since only the DIRECT reference panel was powered to identify any loci for the SIRD, SIDD, MOD, and MARD subtypes. With DIRECT, we found 36 significant associations for SAID, three for SIRD, and one for each of SIDD, MOD, MARD (Supplementary Table 9). Despite whole blood not being the relevant tissue, the discovered loci were frequently directly related to the disease subtype in question: severe autoimmune diabetes was associated with immune genes including *HLA-B-C, LST1*, and *MICA-B;* and *TRMT2A*, a gene implicated in beta-cells apoptosis, was associated with the severe insulin deficient subtype. This demonstrates that a well powered reference panel, even from a non-relevant tissue, can compensate for a lack of power in the original GWAS study to allow us to discover relevant biology.

## Discussion

Our findings show that the relevance of the tissue does not play a large role in the numbers of genes discovered by TWAS methods; instead the sample size of the reference panels was the most important determinant. Not only did we find more genes using the largest available gene expression reference dataset, we also reported a substantial increase in associated genes by using a larger GWAS dataset for T2D than previously published ^31^(from 873 to 1,818 genes). However, we also saw some evidence that our power to discover causal genes using cis-genetic effects may be beginning to become saturated at around the size of the DIRECT reference panel (∼3,000 samples). This sample size is feasible for less accessible tissues by combining datasets; for example recently a consortia has mapped eQTLs using a dataset of 2,970 brain cortex samples from individuals of European ancestry^32^, while others have combined datasets of pancreatic islets to obtain larger sample sizes^5^. It would be interesting to investigate if tissue relevance is more important when using reference datasets of this size, with increased ability to map tissue specific signals.

Comparing the performance of different omics (in this case transcriptomics and antibody based proteomics) to identify mediating molecular phenotypes, we observe that proportionally these assays have comparable power. However, the genome-wide nature of RNA-seq does mean that considerably more hypotheses can be tested using a single assay, while the power of proteomics assays may be boosted by the pre-selection of proteins directly relevant for disease. Though it was not significant due to small numbers, we observed a large odds ratio for enrichment when looking at expression and protein associations. This may be partly due to TWAS methods only considering genetic effects in cis, or local to the gene, and so excludes effects specific to proteins such as post transcriptional modification effects. This also suggests that approaches which look to combine eQTL and pQTL information when identifying gene targets could be reusing the same information ^33^, obtained using the two different assays, which could lead to overestimation of a gene’s suitability as a target. An alternative approach could focus on trans effects on proteins, less likely to be mediated by mRNA.

Type 2 diabetes is a complex disease involving different processes and tissues. Among these processes, insulin resistance is related to obesity and tissues such as skeletal muscle and adipose. By controlling for BMI as a proxy for obesity, we expected to remove genetic signals related to these specific tissues, but instead we observed a uniform decrease across all tissues, including pancreatic islets known to be related to beta-cell dysfunction. One reason could be that the original GWAS mainly included individuals of European ancestry, it is known that the development of T2D in such cohorts is less driven by beta-cell dysfunction than cohorts of other ethnicities^34^. A consequence of this could be that the study had greater power to discover insulin resistance related causes of T2D. However, this may also reflect the fact that beta-cell dysfunction is related to obesity as well, and in particular to the accumulation of liver fat^35^ and that BMI is a poor predictor of T2D in some groups of individuals. To produce a better understanding of particular processes which drive the development of T2D, studying particular populations affected in greater proportion by these forms of T2D constitutes another approach, alongside expanding reference datasets to reliably map tissue specific effects in inaccessible tissues.

This study has discovered a large number of genes across a large number of tissues, in part due to the large sample size in both the original GWAS and the reference panels used. Because T2D is a high heterogeneous disease, involving many tissues and many genes, this means that many of the implicated loci will have low effect size and be of unclear biological function. Studies of T2D have addressed this heterogeneity by constructing subtypes based on clinical phenotypes^29^, and shown that these subtypes have different genetic associations^30^. However, because of the necessity of collecting extra clinical phenotypes on these individuals, GWAS studies into subtypes are more limited in sample size, meaning we were not able to find any causal loci for most of these subtypes using relevant tissue expression data. However, with the increased power of the DIRECT reference we were able to discover loci of function directly relevant to the subtype under investigation. This suggests that a strategy of using carefully defined disease phenotypes with expression data that prioritizes sample size over tissue of relevance can identify candidate loci for specific disease processes.

In this paper we have demonstrated that cis-eQTL information from current bulk RNA-seq datasets are insufficient for answering questions on causal tissues for disease and tissue specific processes, a fact that others have also noted^36^. eQTL studies using single cell RNA-seq data from multiple individuals are beginning to be produced, these may have the potential to identify cell and tissue specific effects and thus inform on tissue specific disease loci^37,38^. However, these studies have lower power than studies in bulk tissue, and so this potential may not be realised in the near future^39^. Concurrently, groups are producing omics data using tens of thousands of participants, with the potential to identify even more disease related loci, in particular in conjunction with smaller GWAS studies with carefully defined phenotypes. Discovery of causal loci and thus gene targets is increasingly driven by synthesising multiple sources of evidence^33^, functional, genetic and molecular, and we have shown here how studies in accessible but not directly disease relevant tissues can be a valuable source of such evidence.

## List of supplementary files

### ST1 Cohort summary

**ST2 Summary of main results in all tissues**. Number of significant associations, before and after p-values correction for multiple testing, for each tissue, percentage of significant associations, total number of genes in the model, relevance of the tissue for T2D, sample size and formatting variables.

**ST3 Summary of T2D known genes occurrences**. 1 for yes 0 for no. Known genes gathered from the output of DisGeNet.

**ST4 Summary of all significant genes occurrences**.

**ST5 Odds ratios of number of genes found nearby significant GWAS variants**. One table per GWAS with odds ratios and corresponding p-value for each tissue.

**ST6 Number of SNPs included in the model being also a significant GWAS variants. ST7 Mendelian randomization summary**.

**ST8 Comparison of results with and without correction for BMI**.

**ST9 Results for analysis on subtypes**. First table on SAID subtype for all tissues and second table summarizing the genes found only for DIRECT in all 5 subtypes.

**ST10-11 Raw results from MetaXcan for each analysis and each cohort**. Output from MetaXcan analyses with and without BMI. Last column indicates the tissue.

**Zenodo link:** 10.5281/zenodo.7957562

## Methods

### Gene model datasets

eQTLs from three studies were used develop models to identify genetically predicted gene expression. The larger study DIRECT (Diabetes Research on Patient Stratification), with 3,029 samples, included 59,972 independent eQTLs from whole blood (https://doi.org/10.5281/zenodo.4475681). This cohort is composed of both pre-diabetics and newly diagnosed T2D patients with blood sample collected from venous blood. All characteristics and analyzed phenotypes are described in a previous paper ^13^. The same study identify 1592 independent pQTLs that were used for protein models. For pancreatic islets models of expression, we used InsPIRE (Integrated Network for Systematic analysis of Pancreatic Islet RNA Expression, n =420), which identify 7741 gene level independent eQTLs after a eQTL analysis using fastQTL (https://zenodo.org/record/3408356). Sample collection, analyses and description of data produced by the consortium has been described in Vinuela et al ^40^. For other tissues, GTEx (Genotype-Tissue Expression) v8 eQTLs were downloaded form the GTEx Portal (https://www.gtexportal.org/home/datasets), including 23,268 independent eQTLs from 49 tissues (73 to 706 samples). All methods from QTL studies to fine-mapping and functional analyses are described in the main paper for this study ^9^.

### GWAS datasets

GWAS summary statistics for T2D from the DIAGRAM study, adjusted and not adjusted for BMI were downloaded (https://diagram-consortium.org/downloads.html), selecting the dataset of European background and the meta-analysis of 32 GWAS with 74,124 cases and 824,006 controls^19^. In addition, we used summary statistics from Spracklen et al, 2020 ^20^ which performed a meta-analysis gathering 23 studies in the 1000 Genomes phase 3 from the Asian Genetic Epidemiology Network (AGEN) consortium (https://blog.nus.edu.sg/agen/summary-statistics/t2d-2020/). Summary statistics were also lifted over to the GRCh build 38 to be analysed in the context of the GTEx reference panels.

### TWAS analysis using MetaXcan

The TWAS main analysis was performed using MetaXcan as described here: https://github.com/hakyimlab/MetaXcan. First, and using eQTL variants IDs and betas as weights we derived the MetaXcan models for gene expression prediction. The covariance between each pair of variants included in each gene-model was then calculated using R base *cov()* function and stored in matrices. Second, using as inputs the models, covariances and GWAS summary statistics we derived associations between genetically predicted gene expression and T2D. Pvalues for associations were corrected for multiple testing using the p.adjust() function in R and the Benjamini-Hochberg method as well as for the rest of the follow-up analyses.

### Enrichment analysis

In order to get the odds ratio of genes found nearby GWAS significant variants, we first selected the list of significant genes for each tissue and extracted the chromosome and positions for them. After doing the same for the GWAS significant variants, we applied the *intersect* command from the bedtools suite^41^ to overlap the two source files in order to filters genes found in a 1MB window around these selected GWAS variants. Then, on R, we used the base odds.ratio() function to get the enrichment of each tissue compared to DIRECT which was selected as the reference for this analysis.

### Identification of T2D known genes

To further investigate if genes identified were known T2D genes we used the DisGeNET platform (https://www.disgenet.org/home/). This online resource is a database of genes and variants associated to diseases publicly available which includes references from GWAS analyses, curated repositories and scientific literature.

### Mendelian Randomization analysis

We used the mr-egger function from the *MendelianRandomization* package in R (https://CRAN.R-project.org/package=MendelianRandomization) which is applying a MR method based on the Egger regression^42^. This method requires for each tested gene the betas and standard errors from the GWAS and the QTL analysis. We tested only significant genes with three or more variants included in the model and estimated MR pvalue and heterogeneity pvalue.

## Data Availability

All data produced in the present study are available upon reasonable request to the authors
Supplementary and raw data also available on Zenodo at : 10.5281/zenodo.7957562

## Authors contributions

David Davtian, T Dupuis^1^, D Mansour Aly^2^, N Atabaki-Pasdar^3^, M Walker^4^, P W Franks^5^, F Rutters^6^, HK

Im^7^, E R Pearson^1^, M van de Bunt^3^, A Viñuela, AA Brown^1*^

Conceptualization: DD, AV, AAB Methodology: DD, HKI, MvdB, AAB Software: DD, TD

Formal analysis: DD, AAB Investigation: DD

Resources: DMA, NAP, MW, PWF, FR, ERP, AV

Data curation: DD, TD, DMA, AV Writing-original draft: DD, AV, AAB

Writing-review & editing: DD, TD, MvdB, AV, AAB Visualization: DD, AAB

Supervision: ERP, AV, AAB, Project administration: ERP, AAB Funding acquisition: AAB

